# Exploring plantar flexor function 12 months after an Achilles tendon rupture

**DOI:** 10.1101/2025.06.03.25328217

**Authors:** Christina Rohde Ruppert, Jesper Bencke, Jens Bojsen-Møller, Kristoffer Weisskirchner Barfod, Morten Tange Kristensen, Per Hölmich, Maria Swennergren Hansen

**Author notes:** Correspondence, Name: Christina Rohde Ruppert, Address: Hvidovre hospital, Kettegård alle 30, 2650 Hvidovre, Denmark.

## Abstract

**Background:** Achilles tendon rupture (ATR) is associated with significant short- and long-term functional deficits, likely in consequence of rupture induced tendon elongation and associated changes in muscle contractile properties.

**Purpose:** To investigate the association between tendon elongation and plantar flexor functional performance during unilateral hopping 12 months post-ATR. Further, the study examined side-to-side performance differences (injured vs. non-injured leg) 6 and 12 months post-ATR.

**Study Design:** Explorative cross-sectional study.

**Methods:** Participants (n=59) were recruited from the cohort of a larger randomized controlled trial. Tendon length was measured using ultrasonography at baseline and at 6 and 12 month post-ATR to determine tendon elongation. Heel-rise height deficit (injured minus non-injured side) and biomechanical variables of unilateral hopping (peak moment (M_peak_), peak concentric and eccentric power (CP_peak_ and EP_peak_) and relative concentric and eccentric ankle joint work (RCW_ankle_ and REW_ankle_)) were measured on both sides at 6 and 12 months post injury.

**Results:** Absolute and relative 12 months post-ATR tendon elongation was 19.3% (CI: 15.8-22.8mm) and 11.2% (CI: 9.2-13.2%) respectively. No significant associations were found between tendon elongation and biomechanical variables or heel-rise height deficit. Although functional improvements were observed between 6 and 12 months, significant deficits in plantar flexor function during hopping persisted at 12 months post-ATR for the injured vs. non-injured leg. Specifically, M_peak_ was 11% lower (p < 0.001), CP_peak_ and EP_peak_ were 36% (p < 0.001) and 28% lower (p = 0.001), respectively, and RCW_ankle_ and REW_ankle_ were 15% (p < 0.001) and 17% lower (p < 0.001), respectively, in the injured leg compared to the non-injured leg.

**Conclusion:** No significant associations were shown between absolute Achilles tendon elongation and plantar flexor functional performance. Furthermore, side-to-side differences were shown to persist at 12 months.

**Level of Evidence: 3:** *Clinical Relevance:* Our study shows that deficits in plantar flexor contractile function persist at 12 months post-ATR and is compensated during hopping by work of muscles that span the knee- and hip-joint. The altered pattern of activation should be taken into consideration during rehabilitation and in return to sport decisions. Further, the present study indicates that heel-rise height has limited value as a proxy for injury induced tendon elongation.

*What is known about the subject:* Large functional differences between the injured and non-injured leg have been observed both in the short- and long-term following ATR. Functional deficits are more pronounced when examined in more demanding tasks such as jogging and jumping.

*What this study adds to existing knowledge:* This study shows functional deficits of the injured leg 12 months post-ATR and thereby confirms what other studies has found. Additionally, the findings indicate that tendon elongation alone does not explain the deficits in plantar flexor function. Further, the findings underline the importance of selecting a test that specifically measures ankle joint function.

## Introduction

The incidence of Achilles tendon rupture (ATR) has increased in recent years ^1,2^, especially in patients with age above 50 years ^1^. Most incidences of ATR occur during sporting activities (>70%) ^2,3^. Although many patients reach satisfactory levels of self-reported physical activity ^4–9^ complete rehabilitation to pre injury level is rarely obtained ^4,6,7,9–16^.

ATR is associated with significant functional deficits, seen as differences in plantar flexor moment, muscle power and joint work between the injured and the non-injured leg, both in the short-^10,16–18^ and long-term ^8,11,13^. Functional deficits are seen regardless of treatment type: surgical or non-surgical ^19–21^ or rehabilitation; standard treatment compared with either early weightbearing ^10,15,16,22^ or late weightbearing ^9^. Studies suggest that functional deficits are potentially related to elongation of the ruptured tendon ^4,6,12,14,23–27^ and concurrent atrophy/shortening of the calf muscles ^14,17,27^ resulting in altered contractile properties of the muscle-tendon unit (MTU). Tendon elongation is shown to occur until 6 months after ATR, and although some tendon shortening is reported hereafter^15,28^, the tendon remains elongated years after ATR ^13,23,29^.

With respect to tendon mechanical properties, the elongation yields a more compliant tendon with a larger loss of energy to hysteresis during rapid, maximal contractile efforts involving a stretch-shortening cycle (SSC) ^23,24^. Therefore, to investigate ankle joint function deficit, it is recommended to use demanding SSC-type tasks ^4,24^ instead of submaximal force exertion activities such as e.g. walking ^7,29,30^. Further, the test should target ankle joint function and therefore knee- and hip dominated tasks like drop jump, countermovement-jump and squat jump may not be optimal tasks of functional assessment following ATR ^4^.

Previous studies have investigated the association between tendon elongation and functional outcomes, but with large variation in procedures and with conflicting results. Some studies reported associations between tendon elongation and ankle joint kinematics ^4,31^, strength outcomes ^32^ or heel-rise work ^6^, while other studies revealed no associations between tendon elongation and strength outcomes ^28^, kinematics ^16,22^ or kinetics ^4,6,33,34^. Although previous reports suggest an association between tendon elongation and reduced ankle joint function ^4,6,26,27,34^, only one study has specifically investigated tendon elongation relative to ankle joint function in a demanding unilateral jumping task ^6^, however, only more generic assessments such as jump height and reactive strength index (flight time/contact time) were reported ^6^.

Therefore, the aim of the present study was to investigate the association between ATR-induced tendon elongation and plantar flexor functional performance during demanding unilateral hopping at 6 and 12 months post-ATR, while functional deficits were assessed by side-to-side differences.

## Methods

The present exploratory study elaborated on previously published data ^7^ that were part of a multicenter Randomized Controlled Trial (RCT) ^35^. The RCT compares the effect of an individualized treatment algorithm: Copenhagen Achilles Rupture Treatment Algorithm (CARTA) vs. control groups of either surgical or non-surgical treatment ^7^. The original RCT was registered at ClinicalTrials.gov the 1^st^ of June 2018 (NCT03543943) and the study protocol has been published^35^. The trial was approved by the National Committee of Health Research Ethics (journal number: 1–10–72–428–17) ^7^. Participants received written and verbal information and signed informed consent. All patient related information was anonymized and stored securely. The study was conducted in accordance with the Declaration of Helsinki ^36^.

The participants in the present study are the same as in Hansen et al. ^7^ and the inclusion/exclusion criteria are detailed in the protocol paper ^35^. A total of 156 patients were screened for eligibility from June 2018 to September 2019. 59 patients were included in the present study (Figure 1). The participants were randomized to different treatments as they were enrolled in the RCT, but the present study did not distinguish between the three RCT treatment groups; CARTA (n = 21), non-surgical (n = 20) or surgical (n = 18) treatment. All patients followed the same rehabilitation program, which is described previously ^35^. In brief, the patients were allowed no-weightbearing during the first three weeks, whereafter partial weightbearing was allowed and progressed. After nine weeks of rehabilitation, patients underwent a home-based exercise program twice a day for four weeks. If additional physiotherapy was required thereafter, patients were referred to rehabilitation in the municipality or a private practice.

**Figure 1:**
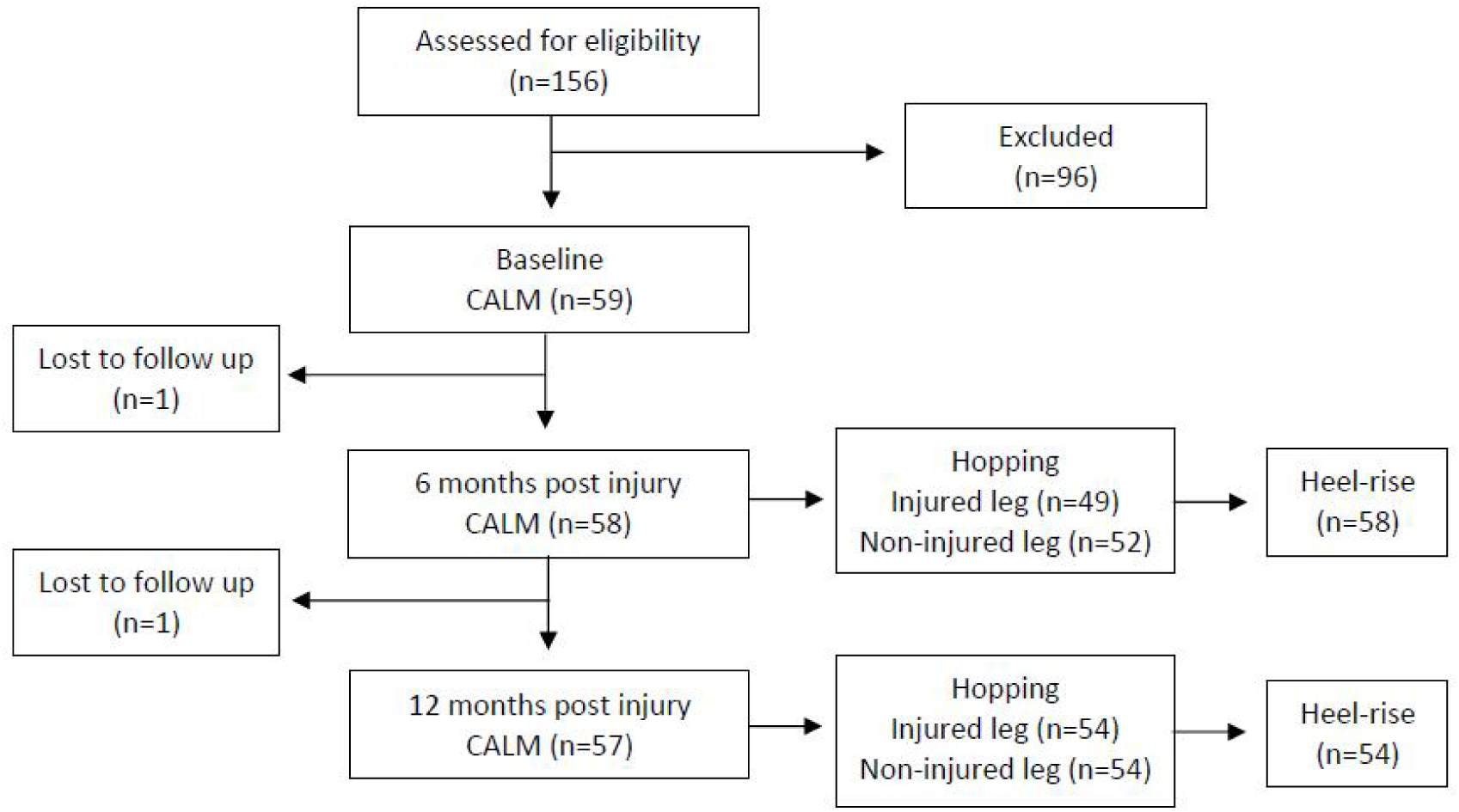
CONSORT flow diagram of the participation from inclusion to 12 months follow-up. Patients were excluded or lost to follow up for various reasons not all specified. 6 months post-ATR some patients were not able to perform the hopping test. 12 months post-ATR some patients were lost for the laboratory testing. Abbreviations figure 1: Copenhagen Achilles Rupture Treatment Algorithm (CARTA), Copenhagen Achilles Length Measure (CALM).

All performance tests were conducted 6 months and 12 months post-ATR at the Human Movement Analysis Laboratory, Copenhagen University Hospital Amager-Hvidovre. Measurements were conducted by physiotherapists who were highly skilled in all the measurements/techniques and familiar with the patient group.

The hopping test was performed as described by Silbernagel et al. ^37^ and has shown both valid and reliable as a measurement for lower leg function in patients with ATR ^26,37,38^. Prior to testing, the patients were familiarized by performing a series of hops in the required frequency (2Hz). Patients were encouraged to maintain their position on the force plate during hopping (Figure 2). In case of failed landings outside the force plate, the affected, the previous and subsequent jump was excluded from post-hoc analysis. The middle 20 hops were averaged for further analysis as previously reported by Silbernagel et al. ^37^. For the biomechanical analyses of repetitive hopping, 22 reflective markers were placed on specific anatomical landmarks, as described by Speedtsberg et al. ^11^ and filmed by 8 T40 infrared cameras (Vicon Motion Systems, Oxford, UK), to create a 3D model for analysis. Simultaneously, ground reaction force data was obtained from two OR6-7 force plates (AMTI) embedded in the floor. Subsequently joint angles, joint moments and power of the ankle, knee and hip joints during hopping were calculated using inherent software of the Nexus (Nexus 2.14.0, Vicon Motion Systems, Oxford, UK). Outcome parameters; peak ankle dorsiflexion (°) (DF_peak_), peak ankle moment (N·m) (M_peak_), peak ankle concentric and eccentric power (W) (respectively CP_peak_ and EP_peak_), absolute ankle, knee and hip concentric and eccentric work (J/kg) and the relative concentric and eccentric phase ankle joint work (%) (respectively RCW_ankle_ and REW_ankle_) were extracted using custom-made MATLAB scripts (MATLAB R2019b).

**Figure 2:**
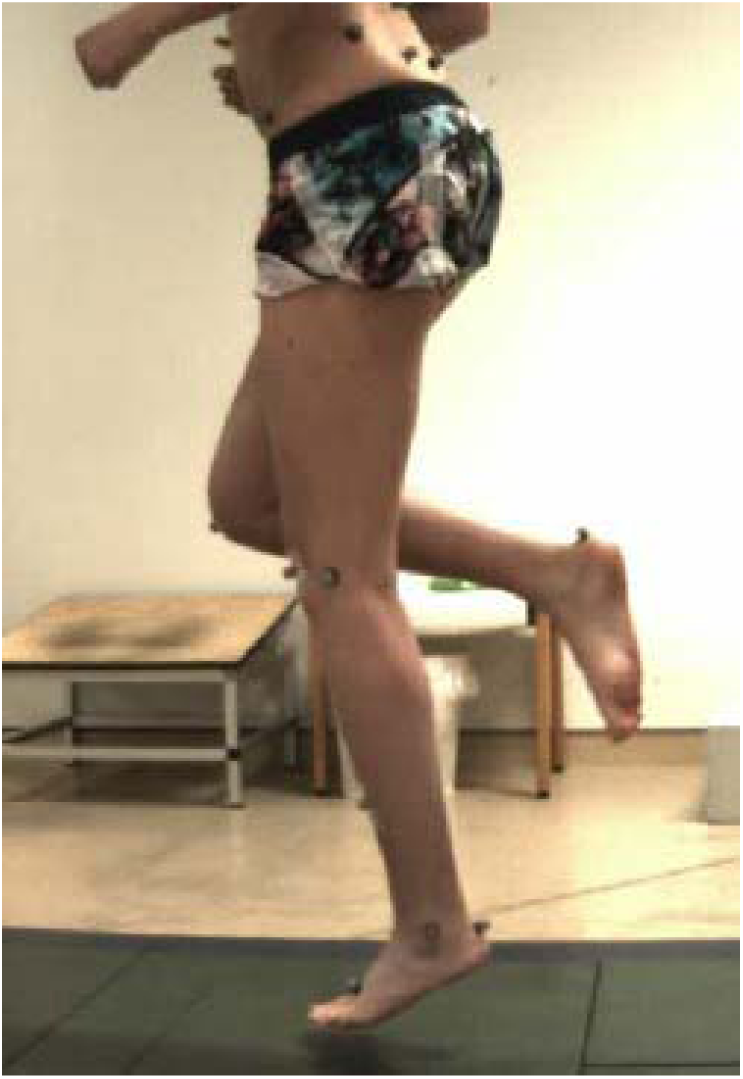
The unilateral hopping test. The unilateral hopping test was performed on a force plate and 22 retroreflective markers placed on anatomical landmarks were filmed by infrared cameras to create a 3D model for analysis.

A limb symmetry index (LSI) was calculated as injured/non-injured leg x 100% for all variables at both 6 and 12 months. At 6 months 49 patients performed the hopping test on both legs while three patients were unable to perform the test on the injured leg. At 12 months 54 patients performed the hopping test bilaterally.

The heel-rise test was conducted as described by Silbernagel et al. ^39^ and has shown to be valid for patients with ATR ^39^. The maximal heel-rise height was used to calculate heel-rise height deficit (injured minus non-injured side).

At baseline, and at 6 and 12 months, measurements of Achilles tendon elongation and patient reported outcomes were conducted. Achilles tendon elongation was quantified by use of ultrasonography applying the Copenhagen Achilles Length Measure (CALM) ^35^. CALM has been shown as a valid ^40,41^ and reliable measurement for Achilles tendon length ^40,42^. Tendon elongation was determined as the absolute (mm) and relative (%) differences in tendon length of the non-injured leg and the injured leg. Patient reported outcomes were evaluated with Achilles tendon total rupture score (ATRS) which is developed and validated for the population ^43^.

All statistical analyses were made in RStudio (v. 2023.12.1-402). The level of significance was set at p<0.05. Demographic data are presented as mean (%95 confidence interval) unless otherwise stated. All data were checked for normal distribution using a Shapiro-Wilks test and by a visualizing histogram. Correlation analyses were performed and presented with Pearson’s R or Spearman’s Rho’s correlation coefficient (p-value) depending on normal distribution. Correlation analysis was made between tendon elongation and heel-rise height deficit at 12 months, and further correlations between tendon elongation or heel-rise height deficit and LSI 12 months post-ATR for selected variables (DF_peak_, M_peak_, CP_peak_, EP_peak_, RCW_ankle_ and REW_ankle_ ) were analyzed. Possible confounding variables were: age, sex, BMI, preinjury level of physical function evaluated with ATRS and currently level of physical function evaluated with ATRS.

Biomechanical variables were analyzed using a 2-by-2 (time (6 months and 12 months) by leg (injured and non-injured)) repeated measures analysis of variance (ANOVA). Data is presented as mean (95% confidence interval) with the result of the ANOVA with the respective p-value.

## Results

Anthropometrics of participants are shown in table 1 for baseline (within four days post-ATR), 6 months and 12 months follow-up. Apart from ATRS, no significant differences were found between the three testing time points.

**Table 1:**
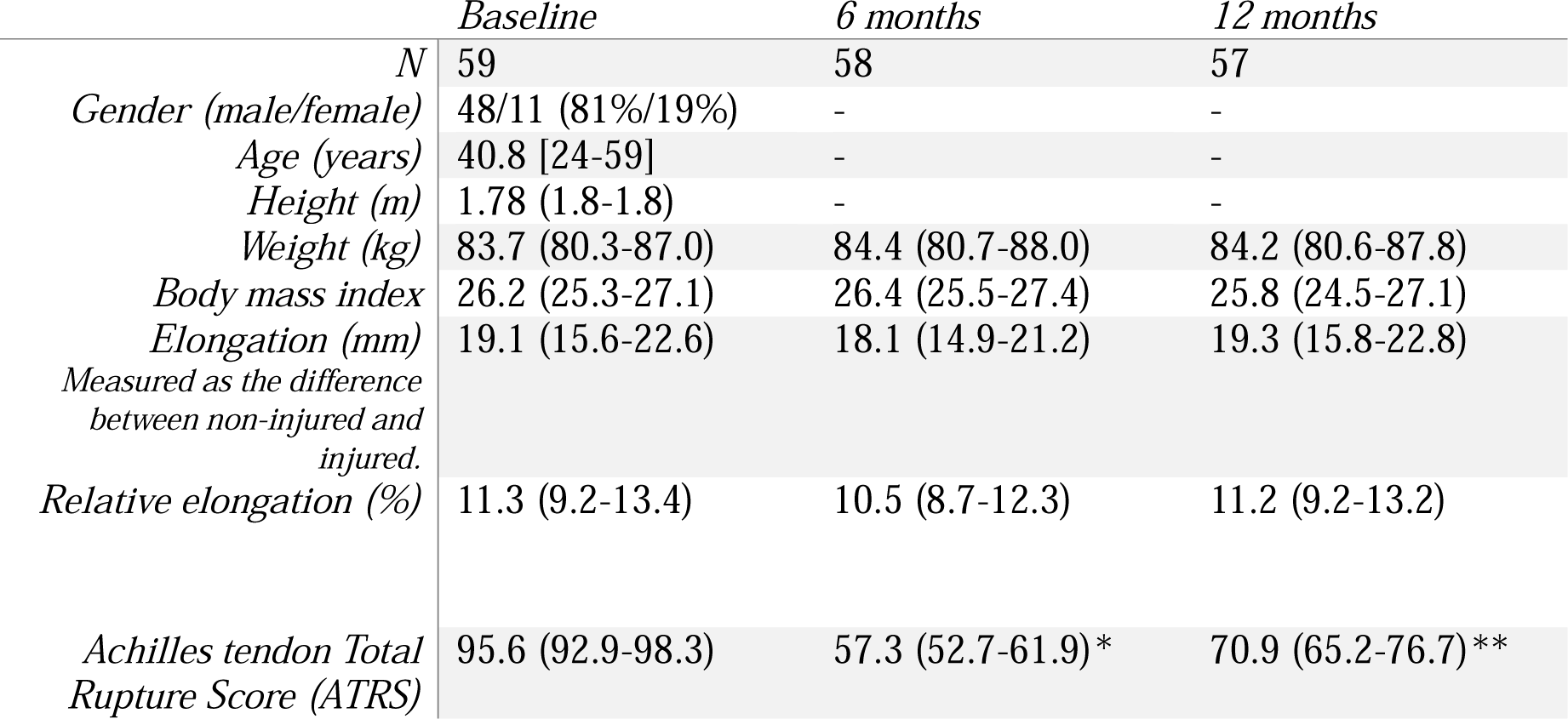
Anthropometic data, Achilles tendon elongation and Achilles tendon Total Rupture Score (ATRS) at baseline and at 6- and 12 months follow up. Data is presented as count (%) or mean where () denotes 95% confidence interval and [] denotes range. *Statistically significant difference between baseline and 6 months. ** Statistically significant difference between both baseline and 12 months and 6 months and 12 months.

A weak but significant negative association was shown between relative tendon elongation and LSI CP_peak_ (Rho = -0.37 p = 0.007). Apart from that, only very weak associations were found between both absolute and relative tendon elongation and functional outcomes of the hopping test (LSI DF_peak_, LSI M_peak_, LSI CP_peak_, LSI EP_peak_, LSI RCW_ankle_, and LSI REW_ankle_) at 12 months (R and Rho values ranged from 0.04 to 0.29, p > 0.055 for absolute elongation, and from -0.26 to 0.25, p > 0.08 for relative tendon elongation). No statically significant association was found between absolute tendon elongation and heel-rise height deficit (injured minus non-injured leg) at 12 months follow-up (Figure 3). However, a weak statistically significant, but not clinically relevant negative association was found between relative tendon elongation and heel-rise height deficit (Rho = -0.38, p = 0.007). Similarly, weak associations were found between heel-rise height deficit and biomechanical outcomes of hopping (Rho values ranged from 0.24 to 0.45, p>0.056, except for RCW_ankle_ (Rho = 0.28, p = 0.05) and CP_peak_ (Rho = 0.45, p = 0.008)). Correlation coefficients, with respective p-values can be found in supplementary material table A.

**Figure 3:**
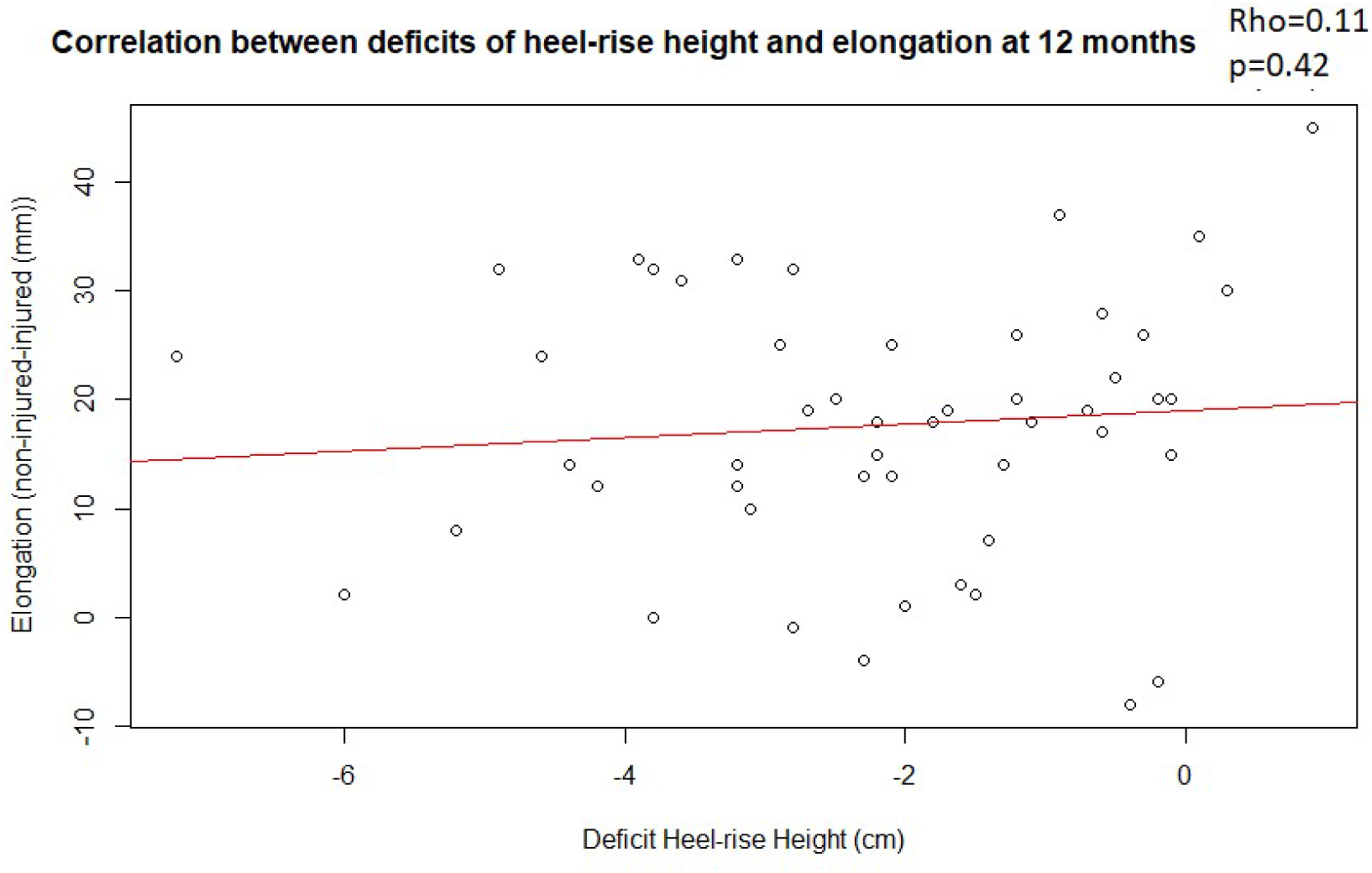
Scatterplot with line of linear regression showing the correlation between heel-rise height deficit and tendon elongation. The correlation analysis shows a very poor association (Rho=0.11, p=0.42) between deficits in heel-rise height and elongation 12 months after Achilles tendon rupture and the scatterplots shows a large spread of the results.

The results of the ANOVA revealed that both time and leg had a statistically significant effect on performance for the variables M_peak_, CP_peak_, EP_peak_, RCW_ankle_ and REW_ankle_ (p < 0.001) (Table 2).

**Table 2:** Results of ANOVA. Mean (95% confidence interval), result of ANOVA (p-value) and the Limb symmetry index (LSI) for 6 and 12 months for the variables; peak dorsiflexion, peak ankle moment, peak ankle concentric and eccentric power, total concentric and eccentric work and relative concentric and eccentric ankle work.

|  | 6 months |  |  | 12 months |  |  | ANOVA |
| --- | --- | --- | --- | --- | --- | --- | --- |
|  | Injured<br>(n=49) | Non-injured<br>(n=52) | LSI (%) | Injured<br>(n=54) | Non-injured<br>(n=54) | LSI (%) |  |
| *Peak dorsiflexion (degree)* | 21.4<br>(20.2-22.5) | 23.6<br>(22.9-24.3) | 90<br>(86-94) | 20.4<br>(20.1-22.7) | 22.8<br>(21.6-24) | 94<br>(88-100) | No interaction<br>between leg and<br>time (p>0.05) |
| *Peak ankle moment (N·m)* | 1.7<br>(1.6-1.9) | 2.4<br>(2.3-2.5) | 73<br>(68-77) | 2.13<br>(2-2.3) | 2.4<br>(2.2-2.5) | 90<br>(87-93) | Main interaction<br>between leg and<br>time (p<0.01) |
| *Peak ankle concentric plantarflexor power (watt)* | 6.2<br>(5.6-6.8) | 10.6<br>(9.8-11.4) | 58<br>(54-62) | 7.81<br>(7.2-8.5) | 10.58<br>(9.8-11.4) | 74<br>(70-78) | Main interaction<br>between leg and<br>time (p<0.01) |
| *Peak ankle eccentric plantarflexor power (watt)* | 5.3<br>(4.7-5.9) | 8.6<br>(7.9-9.3) | 61<br>(56-66) | 6.7<br>(6.1-7.3) | 8.6<br>(8-9.2) | 81<br>(76-85) | Main interaction<br>between leg and<br>time (p<0.01) |
| *Total concentric work (joule/kg)* | 22.7<br>(21.5-23.9) | 27.4<br>(26.2-28.6) | 83<br>(80-87) | 23.2<br>(22-24.4) | 26.2<br>(24.9-27.4) | 89<br>(85-93) | Main effect of<br>leg (p<0.01) |
| *Relative concentric ankle work (%)* | 49.8<br>(47.2-52.4) | 68.8<br>(66.2-71.3) | 73<br>(69-76) | 59<br>(56.5-61.5) | 69.3<br>(66.4-72.2) | 87<br>(83-90) | Main interaction<br>between leg and<br>time (p<0.01) |
| *Total eccentric work (joule/kg)* | 17.7 (16.6-18.7) | 20.7 (19.6-21.7) | 86 (82-91) | 18.7 (17.6-19.8) | 19.8 (18.9-20.8) | 97 (91-103) | Main effect of<br>leg (p<0.01) |
| *Relative eccentric ankle work (%)* | 38<br>(34.6-41.4) | 56.1<br>(53.2-59.1) | 68<br>(63-73) | 47.9<br>(44.6-51.2) | 57.9<br>(55.3-60.6) | 83<br>(78-89) | Main interaction<br>between leg and<br>time (p<0.01) |

Improvements of 24% in LSI M_peak_ were found from 6 to 12-months (p < 0.001) (Figure 4). 12 months post ATR the injured leg demonstrated 11 % lower M_peak_ compared to the non-injured leg (p<0.001). 51% of the patients had LSI M_peak_ of ≥90%.

**Figure 4:**
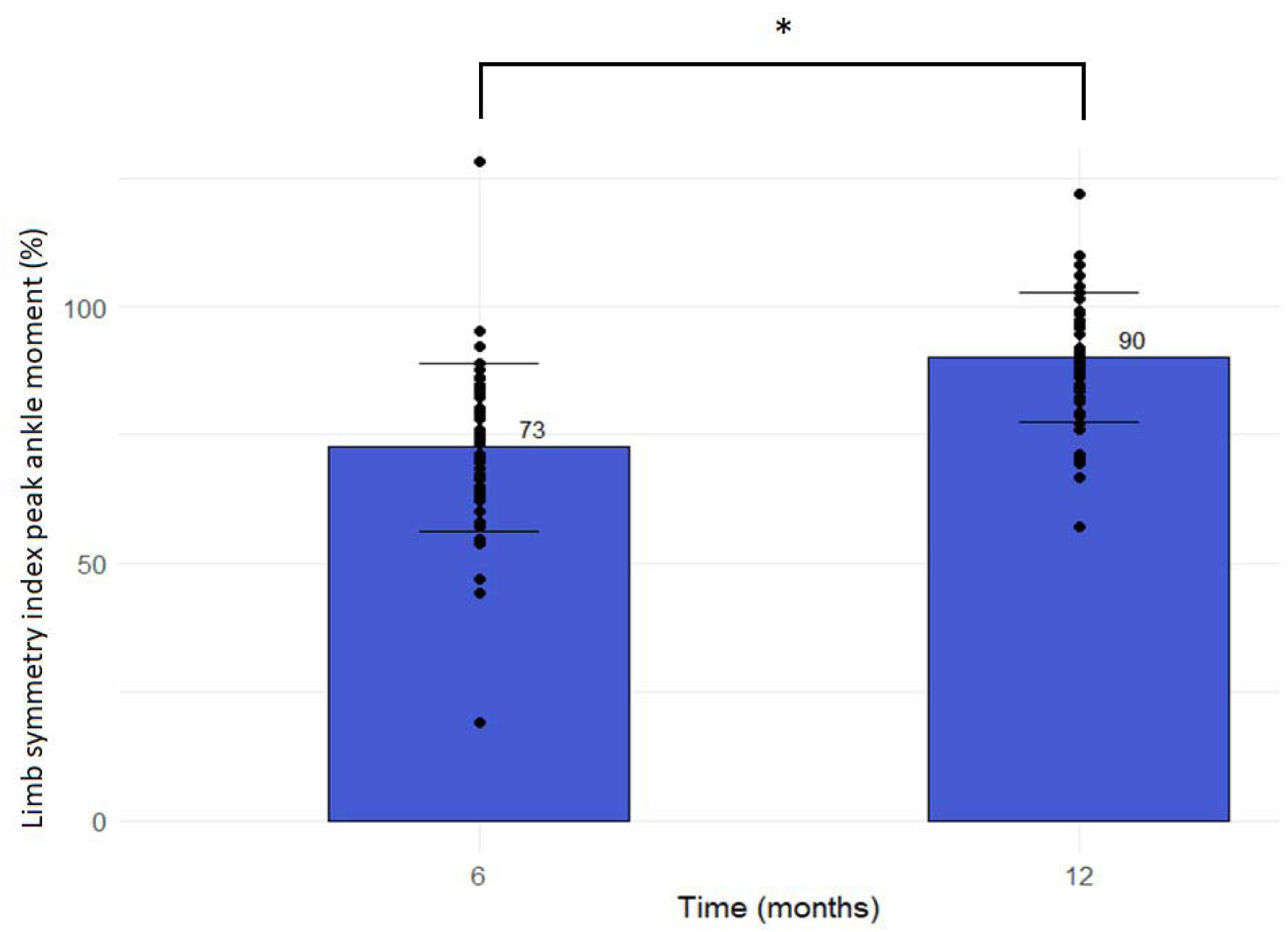
Limb symmetry index (LSI) ankle peak moment 6- and 12 months after Achilles tendon rupture. LSI M_peak_ increased by 24% from 6- to 12-months after ATR (p<0.001).

Patients improved LSI CP_peak_ and EP_peak_ with 29% (p<0.001) and 33% (p<0.001) respectively, from 6- to 12-months (figure 5). 12 months post ATR the injured leg demonstrated 36% (p<0.001) lower CP_peak_ and 28% (p<0.001) lower EP_peak_ compared to the non-injured leg. Further, only 13% of the patients had CP_peak_ LSI ≥90% and only 22% had EP_peak_ LSI ≥90%.

**Figure 5:**
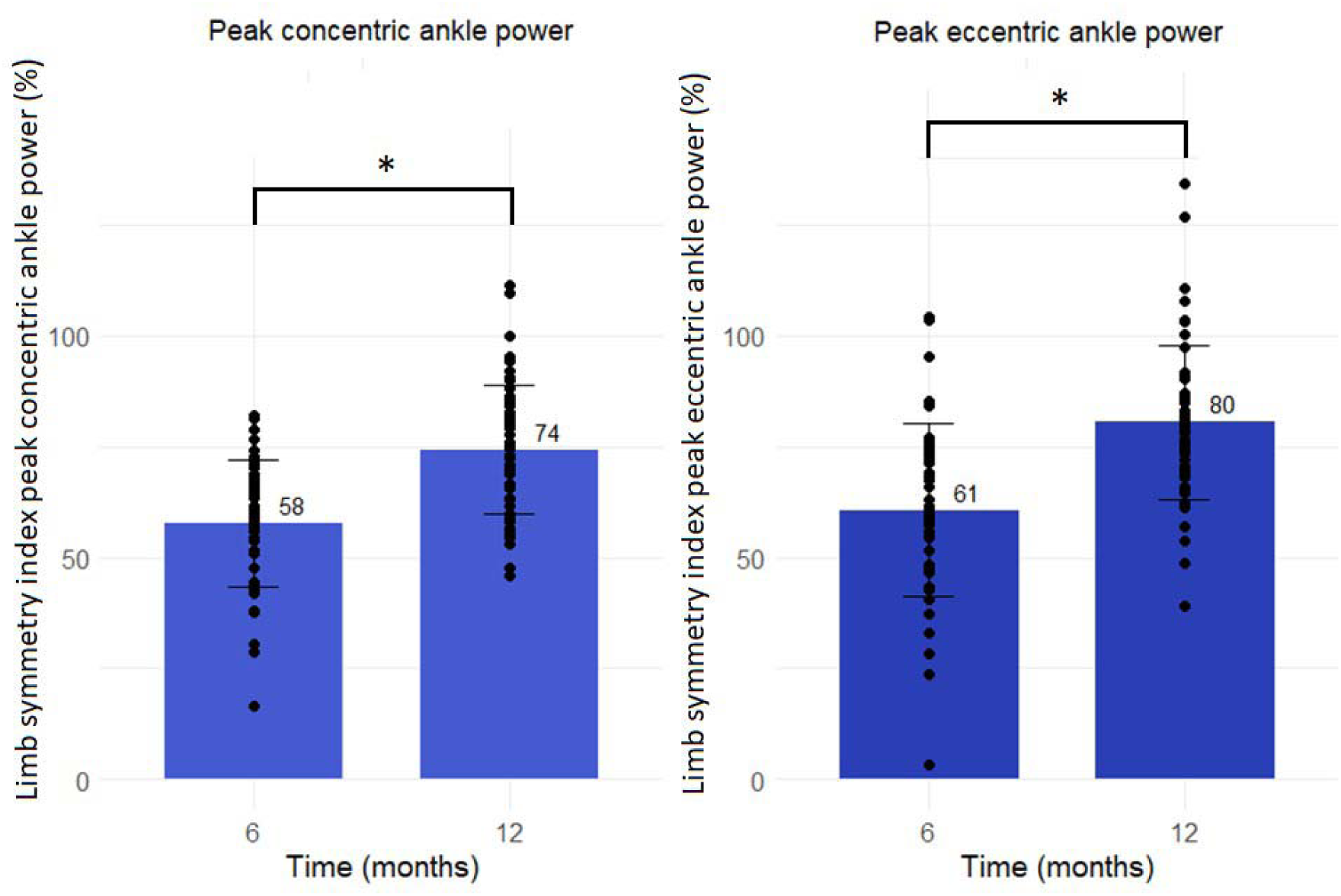
Limb symmetry index (LSI) of ankle concentric (left) and eccentric (right) peak power 6- and 12 months after Achilles tendon rupture. Limb symmetry index concentric peak power increased by 29% (p<0.001) and Limb symmetry index eccentric peak power increased by 33% (p<0.001) from 6 to 12 months after Achilles tendon rupture.

Total work differed significantly between legs, with the injured leg showing 12% lower work in the concentric phase (p < 0.001) and 6% lower in the eccentric phase (p = 0.04) 12 months post-ATR. However, and most importantly, the distribution of work between the ankle, knee and hip joints was different 12 months post-ATR. Also, the injured leg demonstrated 14% lower (p < 0.001) RCW_ankle_ and 17% lower (p < 0.001) REW_ankle_ compared to the non-injured leg 12 months post-ATR (Figure 6). Further, statistically significant increases of relative concentric and eccentric hip joint work were shown for the injured leg (see supplementary material table B).

**Figure 6:**
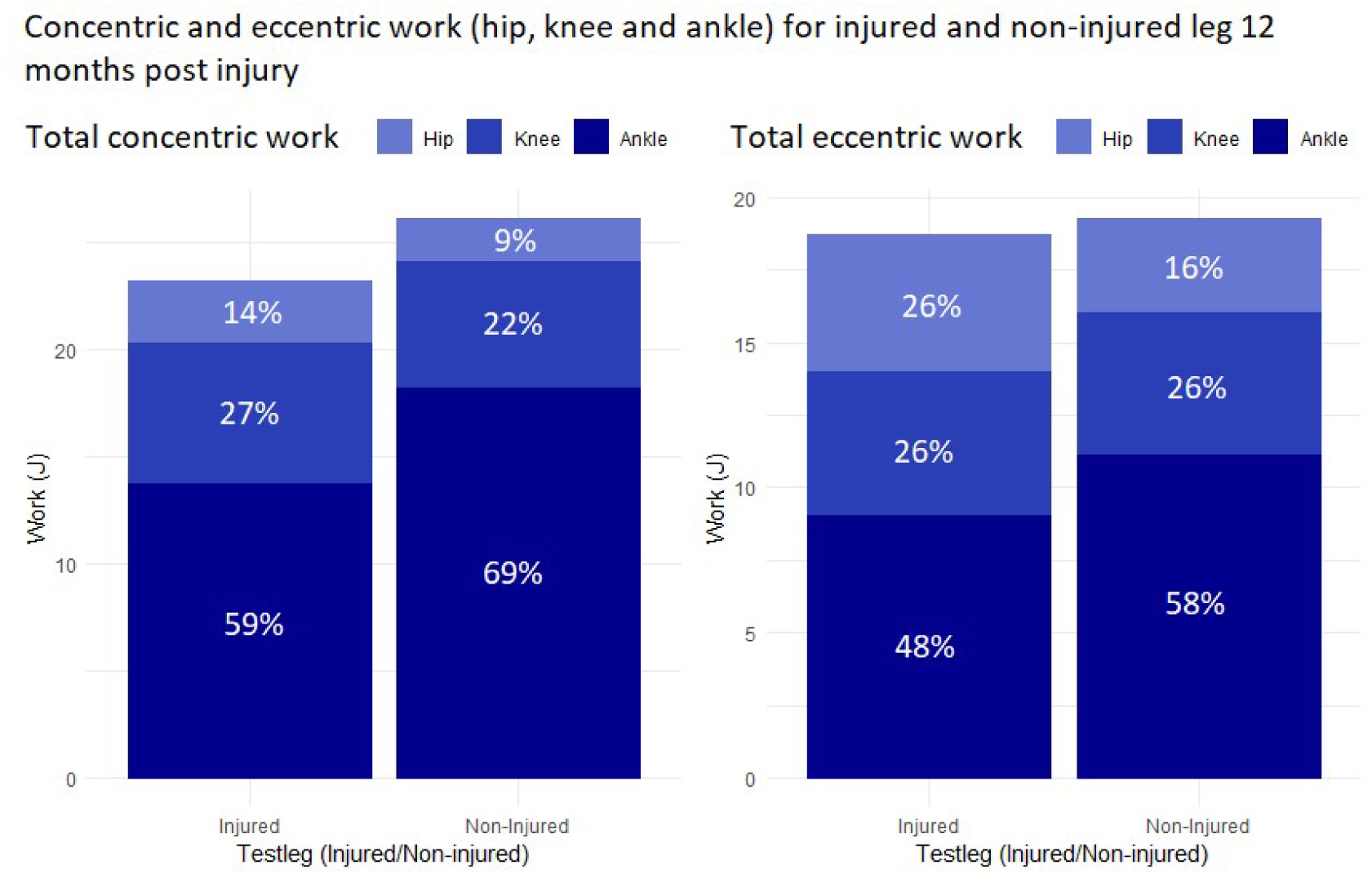
Total concentric- and eccentric work for injured and non-injured leg at 12 months follow-up. The numbers in the columns shows the relative contribution of respectively concentric work (left) and eccentric work (right). 12 months after Achilles tendon rupture the injured leg demonstrated 14% lower (p<0.001) RCWankle and 17% lower (p<0.001) REWankle compared to the non-injured leg.

Adjusted analysis including identified confounders; gender, age, baseline BMI, ATRS pre-injury and ATRS at 12 months follow-up, did not change findings when compared to unadjusted models.

## Discussion

The main findings were that both tendon elongation and heel-rise height deficit showed no significant associations with any of the functional outcomes of the hopping test at 12 months post-ATR, thus indicating that tendon elongation *per se* may not explain deficits in functional performance. Other important findings are that during hopping neither moment, power or work appears to be fully recovered 12 months post-ATR, nor is AT length, which may contribute to altered functional contractile parameters.

ATR has previously been shown to induce tendon elongation, increase tendon compliance and reduce ability to store and release energy during SSC movements ^44^. Therefore, although mechanical tendon properties and hysteresis was not quantified directly in the present study, the lack of association between tendon elongation and functional performance during hopping was unexpected. Thus, there seems not to be consensus in the literature about whether, and to which extent, tendon elongation and functional outcomes are associated. Nonetheless, the inconsistency in the findings might be explained by differences in outcome variables, treatment (surgical or non-surgical), type and/or length of rehabilitation, time of testing and/or in measurement techniques.

Therefore, care should be taken, when comparing studies that evaluate the association between tendon elongation and ankle function after ATR.

Average heel-rise height during a repetitive heel-rise test have been extensively used to evaluate functional outcome after ATR ^4,6,12,16,26,33,39^. Some previous studies have reported negative correlations between heel-rise height deficit and tendon elongation ^4,26,33,45^ and thereby indicated that heel-rise height deficit can be used as a surrogate for tendon elongation. Contrarily, the present data suggest that tendon elongation was weakly associated with heel-rise height deficit, which is in line with other findings ^4,6,33^. Our results suggest that care should be taken to use heel-rise height as a proxy for tendon elongation. It may be speculated that the lack of strong association can be explained in differences in treatment, type and length of rehabilitation or measurements of tendon elongation.

In the present study, kinetic variables were investigated during a demanding unilateral hopping test. Our results show the importance of detailed biomechanical testing, as large differences between injured and non-injured leg were apparent when the concentric and eccentric work was calculated for each joint. Studies evaluating performance as overall lower limb performance (eg. Olsson et al. (8)) may thus lack the joint specific deficiencies related to the injury, as the performance may derive from an increased contribution from the proximal joints. Along this notion the present data indicate that the deficits in concentric and eccentric ankle work following ATR were compensated by an increase in especially hip generated work during hopping. Other studies have similarly shown an increase in knee generated power, but did not report hip joint generated power ^23,24^. Even though the type of biomechanical testing used in the present study is not practically possible in most clinics, these tests are of relevance for the patients and physiotherapists, as they provide detailed evaluations of ankle performance in more demanding tasks and thereby serve to guide the rehabilitation.

Furthermore, our study shows that plantar flexor function remains impaired 12 months post-ATR, consistent with findings from previous research ^4,6,7,9,14,15,26,46^. Some studies have also suggested a larger focus on promoting knee health with preventive strengthening exercises and a lower frequency of demanding exercises including running and jumping for patients with ATR in order to avoid overloading of the knee joint ^14,23,24^. The present study adds additional clinical implications, as it seems that preventive strengthening exercises should target muscles around both the knee and hip joints, especially in the early rehabilitation, which is also suggested by Zellers et al. ^25^.

Moreover, ankle joint training exercises should target both contractile strength and power to the extent possible for the present patient population. However, further studies are needed to investigate the type and timing of training targeting strength and power in patients with ATR.

Although improvements in moment, power, and work were observed between 6 and 12 months, patients still exhibit functional deficits in demanding activities such as hopping 12 months post-ATR. Previously published data on the same patient cohort demonstrate that by 6 months post-ATR, most patients have partially regained their walking ability ^7^. Furthermore, an itemized assessment of the ATRS shows that while patients report a satisfying functional level for daily activities, deficits persist in more demanding activities ^7^ corresponding to the findings of the present study, In the present study, patients were instructed to self-exercise from week 13 after 4 weeks of home-based exercises unless they opted for physiotherapy in the municipality or in a private practice. At this stage, elongation has been shown to occur ^15^ and rehabilitation is suggested to focus on ankle mobilization, proprioceptive and neuromuscular training ^17^. In our experience the majority of the patients had terminated their rehabilitation in the municipality or in a private practice at this time due to satisfactory function of daily activities. However, as the present study demonstrates, plantar flexor function remains insufficient for more demanding tasks. This raises a debate on whether these functional deficits result from inadequate treatment and/or rehabilitation, and whether patients are discharged from supervised physiotherapy too early to ensure full recovery for more demanding activities. Further studies are needed to address these questions.

The present report constitutes a secondary analysis of a larger RCT-study ^35^, therefore the present study includes both surgically and non-surgically treated patients. This might have led to differences in tendon elongation between groups, however as previously reported, no significant differences in tendon elongation between treatments groups (CARTA, surgically or non-surgically) or between surgically and non-surgically treated patients were found ^47^. Therefore, the groups were merged in this study and differences in biomechanical variables between groups were not analyzed in the present study.

Finally, this study is an explorative study and no power analysis has been performed. Therefore, the study is by definition unable to give confirmatory conclusions.

## Conclusion

Despite satisfactory everyday function, patients with ATR are not fully recovered in more demanding movement tasks 12 months after injury. Large side-to-side deficits exist, and the tendon remains elongated at 12 months. However, the present data suggest that other factors than tendon elongation underlie the functional deficits. Compensatory activation strategies (other joints), reduced muscle length or muscle atrophy may play a role for the side specific functional loss, which underscores the importance of whole leg strength and power training for an extended period following ATR.

## Data Availability

All data produced in the present study are stored securely and are not available due to patient confidentiality and data protection regulations

## Acknowledgements

The authors want to thank the staff in the Orthopedic Department, with a special thanks to physiotherapist Julie Jenlar, for contribution with the data collection. Further, the authors want to give a humble thank you to all participants for commitment in the study.

