## supplementary material table A for "Exploring plantar flexor function 12 months after an Achilles tendon rupture"

Table A

Correlations between elongation or heel-rise height deficits and functional outcomes

| *Variable 1* | *Variable 2* | *Correlation Coefficient* | *p-value* |
|---|---|---|---|
| *Elongation* | *Heel-rise height deficit* | Rho=0.11 | p=0.42 |
| *Relative elongation* | *Heel-rise height deficit* | Rho=-0.38 | p=0.006 |
| *Ankle dorsiflexion* | *Elongation* | Rho=0.04 | p=0.75 |
| | *Relative elongation* | Rho=0.25 | p=0.08 |
| | *Heel-rise height deficit* | Rho=-0.24 | p=0.09 |
| *Ankle peak moment* | *Elongation* | R=0.26 | p=0.50 |
| | *Relative elongation* | R=-0.16 | p=0.28 |
| | *Heel-rise height deficit* | Rho=0.26 | p=0.06 |
| *Ankle peak concentric power* | *Elongation* | R=0.12 | p=0.39 |
| | *Relative elongation* | R=-0.37 | p=0.007 |
| | *Heel-rise height deficit* | Rho=0.45 | p=0.0008 |
| *Ankle peak eccentric power* | *Elongation* | Rho=0.25 | p=0.09 |
| | *Relative elongation* | Rho=-0.08 | p=0.56 |
| | *Heel-rise height deficit* | Rho=0.27 | p=0.05 |
| *Relative concentric ankle work* | *Elongation* | R=0.29 | p=0.52 |
| | *Relative elongation* | R=-0.16 | p=0.26 |
| | *Heel-rise height deficit* | Rho=0.277 | p=0.05 |
| *Relative eccentric ankle work* | *Elongation* | Rho=0.27 | p=0.06 |
| | *Relative elongation* | Rho=-0.04 | p=0.80 |
| | *Heel-rise height deficit* | Rho=0.27 | p=0.06 |

*Table A: shows correlation between absolute Achilles tendon elongation or relative Achilles tendon elongation and heel-rise height deficit 12 months after ATR, and between either absolute Achilles tendon elongation, relative Achilles tendon elongation or heel-rise height deficit and variables of LSI 12 months after ATR. Only poor agreement was shown with R or Rho values below 0.5.*
