## supplementary material table B for "Exploring plantar flexor function 12 months after an Achilles tendon rupture"

Table B

Contribution of ankle, knee and hip joint work at 6- and 12 months post ATR

| *Work (% of total work)* | *Joint* | *Leg* | *6 months* | *12 months* |
|---|---|---|---|---|
| *Concentric* | Ankle | Injured | 49.8 (47.2-52.4) | 59.0 (56.5-61.5) |
| | | Non-injured | 68.7 (66.1-71.2) | 69.3 (66.3-72.2) |
| | Knee | Injured | 30.6 (28.1-33.2) | 27.5 (25.4-29.5) |
| | | Non-injured | 22.2 (20.2-24.3) | 22.1 (20.2-23.9) |
| | Hip | Injured | 19.6 (16.9-22.2) | 13.6 (10.6-16.6) |
| | | Non-injured | 9.1 (7.4-10.8) | 8.7 (5.8-11.6) |
| *Eccentric* | Ankle | Injured | 38.0 (34.5-41.4) | 47.0 (44.6-51.3) |
| | | Non-injured | 56.1 (53.2-59.0) | 57.9 (55.3-60.6) |
| | Knee | Injured | 28.8 (26.2-31.4) | 26.3 (24.5-28.9) |
| | | Non-injured | 26.7 (24.5-27.6) | 25.8 (24.0-27.6) |
| | Hip | Injured | 33.3 (30.5-36.0) | 25.9 (22.6-29.1) |
| | | Non-injured | 17.2 (15.3-19.0) | 16.3 (14.5-18.0) |

*Table B: Mean (95% confidence interval) of the contribution of total work between joints (ankle, knee and hip) for concentric and eccentric work for injured and non-injured leg 6- and 12 months after ATR.*
